# Determining Cost-Saving Risk Thresholds for Statin Use

**DOI:** 10.1101/2024.08.14.24311999

**Authors:** Afschin Gandjour

**Author notes:** Corresponding author: Prof. Afschin Gandjour, Frankfurt School of Finance & Management, Adickesallee 32-34, 60322 Frankfurt am Main, Germany.

## Abstract

**Background:** Statins are widely prescribed to reduce cholesterol levels and decrease the risk of cardiovascular diseases. This study aims to determine cost-saving risk thresholds for statin use in Germany to inform better prescription practices and healthcare policies.

**Methods:** A decision-analytic approach was employed, using secondary data to compare statin use with no statin use from the perspective of German sickness fund insurees. The analysis focused on cost savings from avoided cardiovascular (CV) events, translating these avoided events into net savings after accounting for treatment costs and potential side effects. The study considered the German adult population insured by sickness funds and used a lifetime horizon for the analysis.

**Results:** The maximum number needed to treat (NNT) to achieve cost savings over 10 years was found to be 40, leading to a minimum risk threshold for savings of 9.8%. It was estimated that approximately 22% of the adult population in Germany has a 10-year CV risk of ≥9.8%, potentially avoiding between 307,049 and 705,537 CV events over 10 years, with net population savings of approximately €18 billion.

**Conclusions:** The current official threshold for statin prescription in Germany, set at a 20% 10-year risk, is too stringent. Lowering the threshold to 9.8% could significantly increase the number of patients benefiting from statin therapy, reducing CV events and generating substantial cost savings. These findings suggest that adjustments to prescription guidelines could improve cardiovascular outcomes and economic efficiency within the German healthcare system.

## Introduction

According to their label, statins should be prescribed alongside a diet and other nonpharmacological measures when these alone do not sufficiently lower cholesterol levels. In Germany, approximately 4 to 6 million people use statins to reduce their cholesterol levels and decrease the risk of cardiovascular diseases (Laufs, 2015). Adherence to statins in Germany is reported at 84% in two recent studies (Salam, 2023; Koenig, 2024). Despite this, a significant number of patients who could benefit from statin therapy remain untreated (Scheidt-Nave, 2013; Knopf, 2017). Internationally, Germany ranked only 25th out of 91 countries in statin utilization among individuals over 40 years old as of mid-2020 (Guadamuz, 2022).

Overuse of statins may also be a concern, particularly in individuals with a low baseline risk of cardiovascular disease, where the absolute benefit of risk reduction is minimal. For these individuals, the potential for side effects may outweigh the small potential benefit. Overuse can also occur when prescribing practices do not align with clinical guidelines, which often recommend statins primarily for individuals with specific risk profiles. Statin intolerance is not negligible and was observed in 12.5% of participants in a German study, though these results carry some uncertainty due to the uncontrolled nature of the study (Parhofer, 2023).

The German Federal Joint Committee (G-BA) has specified that outside of secondary prevention, lipid-lowering drugs can only be prescribed for patients with a high cardiovascular risk (over 20% event rate/10 years based on available risk calculators). This restriction is justified because, except in cases of secondary prevention, lifestyle changes (e.g., weight reduction and dietary measures) are the first-line therapy for hyperlipidemia (G-BA, 2020).

In 2021, the number of hospital admissions with ischemic stroke in Germany was 216,923 (Ungerer, 2024). Approximately 80% of strokes are ischemic (Wikipedia, 2024). Additionally, around 280,000 people in Germany suffer a heart attack each year (Wikipedia, 2024). A recent commentary by Baldus and Lauterbach (2023) emphasizes that Germany can learn from other developed countries in tackling cardiovascular morbidity and mortality. Primary prevention of cardiovascular disease will remain crucial, requiring systematic identification and control of cardiovascular risk factors.

Statin trials suggest that reducing LDL cholesterol with statins lowers the risk of major vascular events, largely irrespective of age, sex, baseline LDL cholesterol, or previous vascular disease, and reduces vascular and all-cause mortality (Cholesterol Treatment Trialists’ (CTT) Collaborators, 2012). Consequently, the number needed to treat is lower in higher-risk groups, making statins more cost-effective in these groups due to the greater number of prevented events per 100 treated patients (Heller, 2017).

While several cost-effectiveness analyses on statin use for preventing cardiovascular disease in Germany have been conducted, mostly before statins became generic (Obermann, 1997; Lauterbach, 2005), there is a lack of information on the current cost savings potential of statins. This study aims to fill this gap by providing updated insights into the economic benefits of statins, thus informing better prescription practices and healthcare policies. Specifically, this study aims to determine the risk threshold for statin prescription in Germany based on predicted cost savings for sickness funds, which insure about 87% of the German population (Vdek 2024).

## Methods

### Conceptual approach

The analysis was conducted from the perspective of German sickness fund insurees. This perspective is relevant for decisions by the Federal Joint Committee, which assesses the annual treatment costs of new innovative drugs. Unlike the perspective of sickness funds, this specific viewpoint includes copayments made by the insurees.

The study considered the German adult population insured by sickness funds, comparing statin use to no statin use.

### Methodological approach

The study employed a decision-analytic approach using secondary data. It considered the health benefits of statin use in terms of avoided cardiovascular (CV) events and translated these avoided events into cost savings, thereby representing a cost analysis.

The measure of benefits was the net savings from avoided CV events, accounting for healthcare costs during additional life years. The harms of statin use were also considered in terms of costs. The analysis was conducted over a lifetime horizon based on the underlying data sources (see Cost Analysis).

The study determined the threshold for the 10-year risk of CV events based on the point of cost neutrality, at which interventions costs equal cost offsets.

The maximum number needed to treat (NNT) to achieve savings over 10 years was calculated as follows:

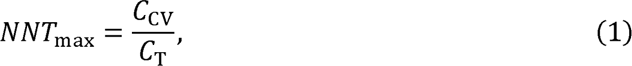

where C_CV_ is the weighted average of the lifetime costs of strokes and myocardial infarctions (MIs), and C_T_ is the treatment cost over 10 years. The formula for C_CV_ is:

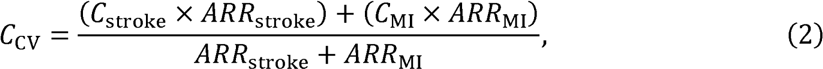

where C_stroke_ is the lifetime cost of stroke, C_MI_ is the lifetime cost of MI, ARR_stroke_ is the absolute risk reduction for stroke, and ARR_stroke_ is the absolute risk reduction for MI.

Next, the minimum absolute risk reduction (ARR_min_) required to achieve the maximum NNT for cost savings was calculated, followed by the minimum baseline 10-year risk for CV events (P_min_) using the relative risk reduction (RRR) of statins:

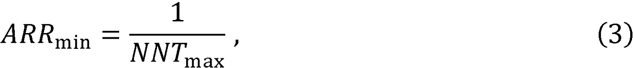

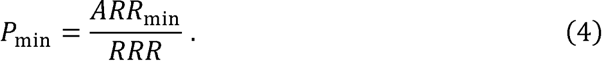

From the risk distribution in the German population, the fraction with a risk above P_min_ was determined. While P_min_ defines the minimum threshold for cost savings, the average risk in the population at or above P_min_ is actually higher. Considering the average risk (P_avg_) is necessary to calculate the number of avoided CV events in the population at or above P_min_. This was determined by subtracting expected CV events with the relative risk reduction from the initial expected CV events based on P_avg_:

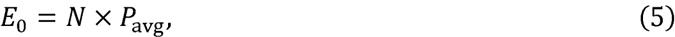

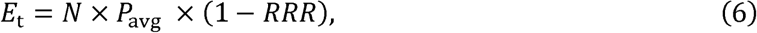

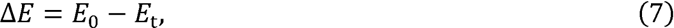

where E_t_ and E_O_ are the number of expected CV events with and without treatment, respectively, and N is the total number of individuals in the population segment.

### Epidemiological data

Based on the meta-analysis by Byrne et al. (2022), a combined relative risk reduction (RRR) for both stroke and myocardial infarction (MI) was calculated using a weighted average approach. This involves weighting each RRR by its respective absolute risk reduction (ARR) and summing these weighted values. The combined RRR for stroke and MI, considering their respective ARRs, is approximately 25.57%. This result is corroborated by a recent systematic review of RCTs showing a median reduction of major CV events by 26% (Dugré, 2023). Most clinical trials report results based on an intention-to-treat (ITT) analysis, which includes all participants as originally allocated after randomization, regardless of their adherence to the treatment regimen. Therefore, the reported RRR typically includes some level of nonadherence.

Using data from the German Health Examination Survey for Adults (DEGS1), we estimated the average 10-year CV risk for individuals in Germany. The DEGS1 survey categorizes individuals into low (<1%), moderate (1-<5%), and high-risk (≥5%) groups based on the SCORE system for CV disease mortality risk. The prevalence of low, moderate, and high risk was 42.8%, 38.5%, and 18.8% in men and 73.7%, 18.1%, and 8.2% in women, respectively (Diederichs, 2018). The SCORE model and related literature often suggest that the incidence of non-fatal CV disease events is approximately 3-5 times higher than that of fatal events.

### Cost analysis

For the cost of statins, the analysis considered the pharmacy retail price after legally mandated prevention of CV diseases (Parhofer, 2023), with 40 mg being the most commonly recommended dose for adults, especially for the prevention of CV events. In a sensitivity analysis, the cheapest statin dose was assumed.

The annual total costs for the treatment and monitoring of a patient taking statins for the prevention of CV diseases range from €31 to €62, excluding medication costs. These costs include regular doctor visits for general and extended examinations (EBM codes 03220 and 03230), costing approximately €15 per visit, occurring 1-2 times per year. Laboratory tests such as lipid profiles (EBM codes 32060, 32061, 32062, 32063) cost €0.15 per test, performed 1-2 times per year, and liver function tests (EBM code 32070) cost €0.25 per test, conducted as needed. Additional specific tests like creatine kinase determination (EBM code 32074) can cost €0.25 per test and are required as needed (Mach, 2020, p. 162).

It was estimated that 12% of statin users in Germany require additional clinical visits annually due to side effects (based on (Parhofer, 2023), with the cost per patient ranging from €19 to €60 based on the type and severity of the side effects. These additional visits are due to issues such as severe muscle pain, new-onset diabetes, significant liver enzyme elevation, and severe gastrointestinal problems.

The data on savings was derived from the lifetime cost of ischemic stroke survivors in Germany, including nursing care, published by Kolominsky-Rabas et al. (2006), based on medical records and interviews. Costs were inflated to 2024 euros, accounting for inflation. The study also considered the increase in health expenditures for treating strokes due to technological advancements, reflecting the decrease in case fatality rates (CFR) of strokes in Germany over time (Rücker, 2020). Improvements in stroke management and treatment, such as increased stroke units and uptake of acute therapies like thrombolytic therapy, contributed to this decrease. The study estimated the cost impact of higher long-term care costs for stroke survivors due to reduced CFR, including rehabilitation costs, outpatient services, and longterm care facilities. Assuming these advancements have added approximately 35% to the costs over 20 years, the estimated inflated expenditure for stroke care in 2024, considering inflation and technological advancements, is approximately €90,245.

This estimate was confirmed by a recently published long-term cost analysis using the InGef (Institute of Applied Health Research) research database with sickness fund claims data for the period from 2014 to 2018 (IGES, 2023). The study reported costs in the first year after an ischemic stroke of €22,404, in addition to a matched control group. After the 6th quarter, insured individuals with an incident ischemic stroke incur approximately €700 per quarter in additional medical treatment costs compared to the control group. Incorporating the control group’s cost of approximately €5,000 per year, the discounted remaining life expectancy of 5.9 years reported by Kolominsky-Rabas et al. (2006), and inflation from 2018 to 2024 yields costs of survivors of about €84,000. It is important to note that discounting future costs in addition to using the discounted remaining life expectancy results in double discounting and is therefore inappropriate. As the analysis has not been peer-reviewed and is limited to a 4-year observational period, its estimate was considered more uncertain and was thus considered the lower limit in a sensitivity analysis.

It would have been possible to consider only the costs of ischemic strokes on top of those of the control group as a saving. However, this approach would have required considering general health expenditures (those of the control group) in added years of survival, necessitating The lifetime cost of MI was predicted to be lower than that of ischemic stroke due to several factors: lower initial costs related to acute care, hospitalization, and follow-up treatments, and lower long-term costs such as medication, lifestyle management, and monitoring. This is because MI typically involves shorter rehabilitation periods and lower rates of long-term disability (Brüggenjürgen, 2005). The study conservatively estimated that the lifetime cost of MI might range between 60% to 80% of the lifetime cost of ischemic stroke. This range was applied in a sensitivity analysis.

The relative risk reduction (RRR) for all-cause mortality is about one-third of that for CV events (Dugré, 2023). Consequently, about two-thirds of individuals who avoid CV events due to statin use would not have experienced a shortened life expectancy due to those events, but they avoid potential health complications and morbidity associated with such events. Based on the simplifying assumption of an exponential survival model, approximately onethird of cost savings from statins would thus be offset by life-prolonging costs, both related and unrelated to CV disease (Gandjour, 2021).

This may present an overestimation of savings because, in patients with CV events (MIs and strokes), not all healthcare expenditures are driven by CV events—approximately 80% of them are (Sidelnikov, 2022). Conversely, it may present an underestimation of savings because, in populations at higher risk (such as older adults or those with existing CV conditions), the hazard rate generally increases over time. The finding that only a portion of cost savings from statins would be offset by life-prolonging costs is consistent with the results of a modeling study using Dutch data. This study suggested that preventing “diseases of the circulatory system” would increase life expectancy while reducing healthcare spending, indicating that savings from avoiding these diseases would not be entirely offset by life extension costs (Grootjans-van Kampen, 2014). In a sensitivity analysis, the fraction of offsets was varied between 20% and 50%. For a discussion of life extension costs associated with statin therapy from various health economic perspectives in Germany, see Wendland et al. (2001).

Discounting was not directly applied as the estimates of the lifetime costs of CV events already incorporated discounting (Kolominsky-Rabas, 2006).

### Sensitivity analysis

In deterministic one-way analyses, uncertainty in the minimum CV risk was assessed by varying individual input parameters susceptible to variation, one at a time, using the boundaries described in Table 1. Furthermore, scenario analysis was provided for the number of CV events avoided and population/individual costs saved (Tables 2 and 3).

**Table 1.** Base-case values and ranges. All costs are in euros.

| Variable | Mean (range) | Reference |
| --- | --- | --- |
| Cost data |  |  |
| Cost of a statin per day | 0.19 (0.12 – 0.19) | Gemeinsamer Bundesausschuss 2024, Bundesministeriums für Gesundheit 2024 |
| Ancillary costs (doctor visits, lab tests) | 47 (31 – 62) | Mach 2020, KBV 2024 |
| Costs of side effects per year | 40 (19 – 60) | Estimate |
| Lifetime cost for stroke care | 90,245 (72,780 – 113,193) | Kolominsky-Rabas 2006 |
| Lifetime cost for MI care | 63,172 (54,147 – 72,196) | Estimate |
| Fraction of cost savings offset by life-prolonging costs | 0.33 (0.20 – 0.50) | Gandjour 2021 |
| Epidemiological data |  |  |
| German adult population with social health insurance | 60,500,000 | Bundesministerium für Wohnen, Stadtentwicklung und Bauwesen 2021, Vdek 2024 |
| Reduction in the number of non-fatal MIs compared to non-fatal strokes | 3.25 (1.92 – 4.58) | Byrne 2022 |
| Relative risk reduction of CV events | 0.25 (0.20 – 0.30) | Byrne 2022 |
| Prevalence of 10-year CV disease risk ≥ 9.8% | 0.22 (0.20 – 0.25) | Diederichs 2018 |

MI: myocardial infarction; CV: cardiovascular

**Table 2.**
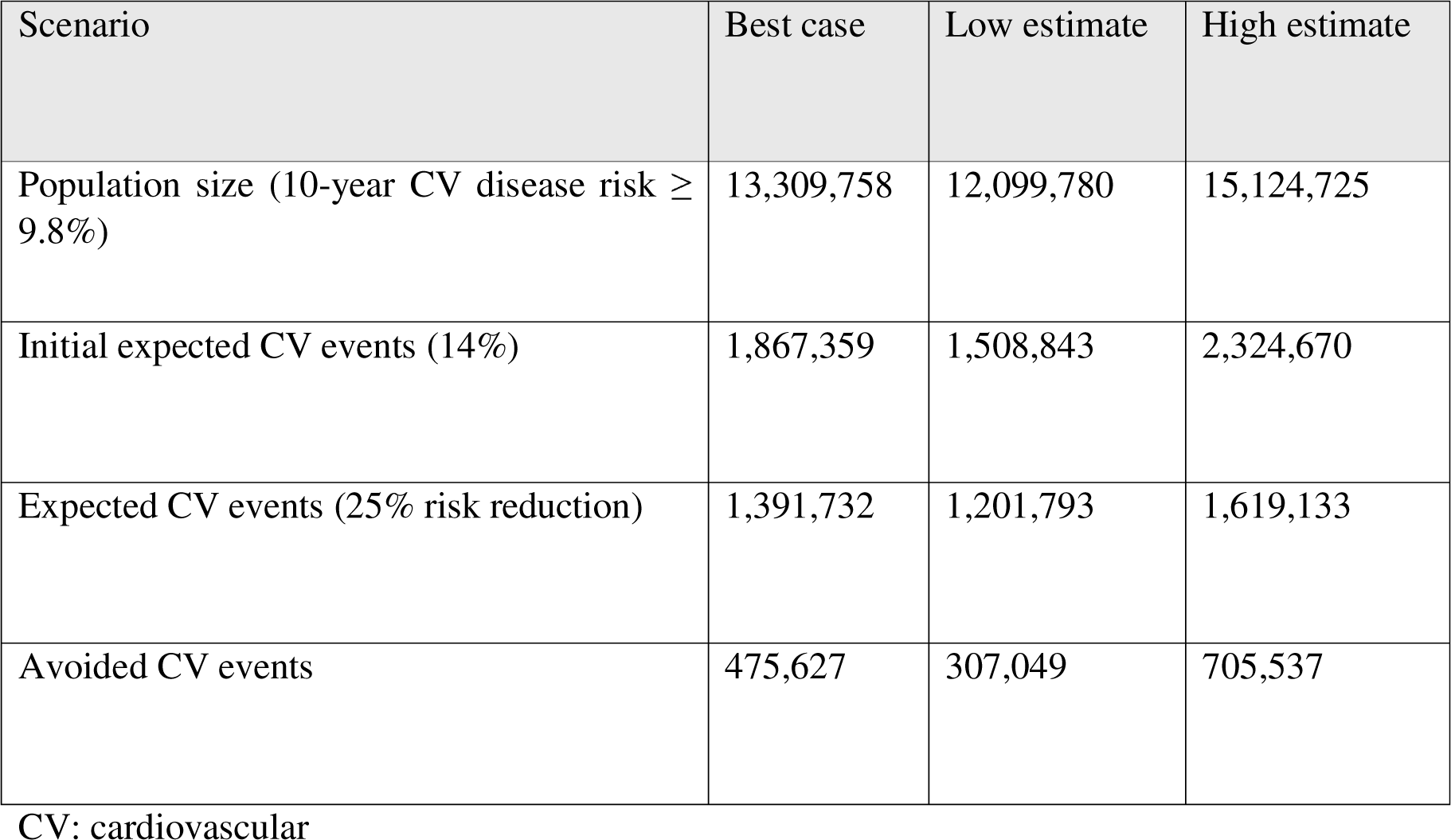
Avoided cardiovascular events over 10 years in sickness fund insurees.

| Scenario | Best case | Low estimate | High estimate |
| --- | --- | --- | --- |
| Population size (10-year CV disease risk ≥ 9.8%) | 13,309,758 | 12,099,780 | 15,124,725 |
| Initial expected CV events (14%) | 1,867,359 | 1,508,843 | 2,324,670 |
| Expected CV events (25% risk reduction) | 1,391,732 | 1,201,793 | 1,619,133 |
| Avoided CV events | 475,627 | 307,049 | 705,537 |

CV: cardiovascular

**Table 3.** Population and individual savings from statin use in the population above the risk threshold. All costs are in euros.

| Scenario | Best case | Low estimate | High estimate |
| --- | --- | --- | --- |
| Intervention cost | 15,389,208,041 | 13,990,189,128 | 17,487,736,410 |
| Savings | 33,075,950,148 | 21,352,752,421 | 49,064,296,193 |
| Net population saving | 17,686,742,107 | 7,362,563,293 | 31,576,559,783 |
| Net individual saving | 1,329 | 608 | 2,088 |

## Results

The maximum NNT reaching the cost-saving threshold was 40, reflecting the ratio of the weighted-average lifetime cost of CV events to the 10-year cost of statin therapy (€46,361/€1156). The resulting minimum risk threshold for savings (P_min_ in Equation 4) was 9.8%, calculated as the ratio of the ARR_min_ to the RRR (2.5%/25%).

Using data from the German Health Examination Survey for Adults (DEGS1), we estimated the average 10-year CV risk for individuals in Germany with a minimum risk of 9.8%. Given that more than 50% of the population falls into the low-risk category (Diederichs, 2018), we focused on the moderate and high-risk groups. Using a log-normal distribution to model the right-skewed nature of CV risk, we calculated an average risk of approximately 14.03% for individuals above the 9.8% threshold (see Figure 2). This aligns well with the DEGS1 data, suggesting that our model accurately reflects the distribution of CV risk in the German population.

**Figure 1.**
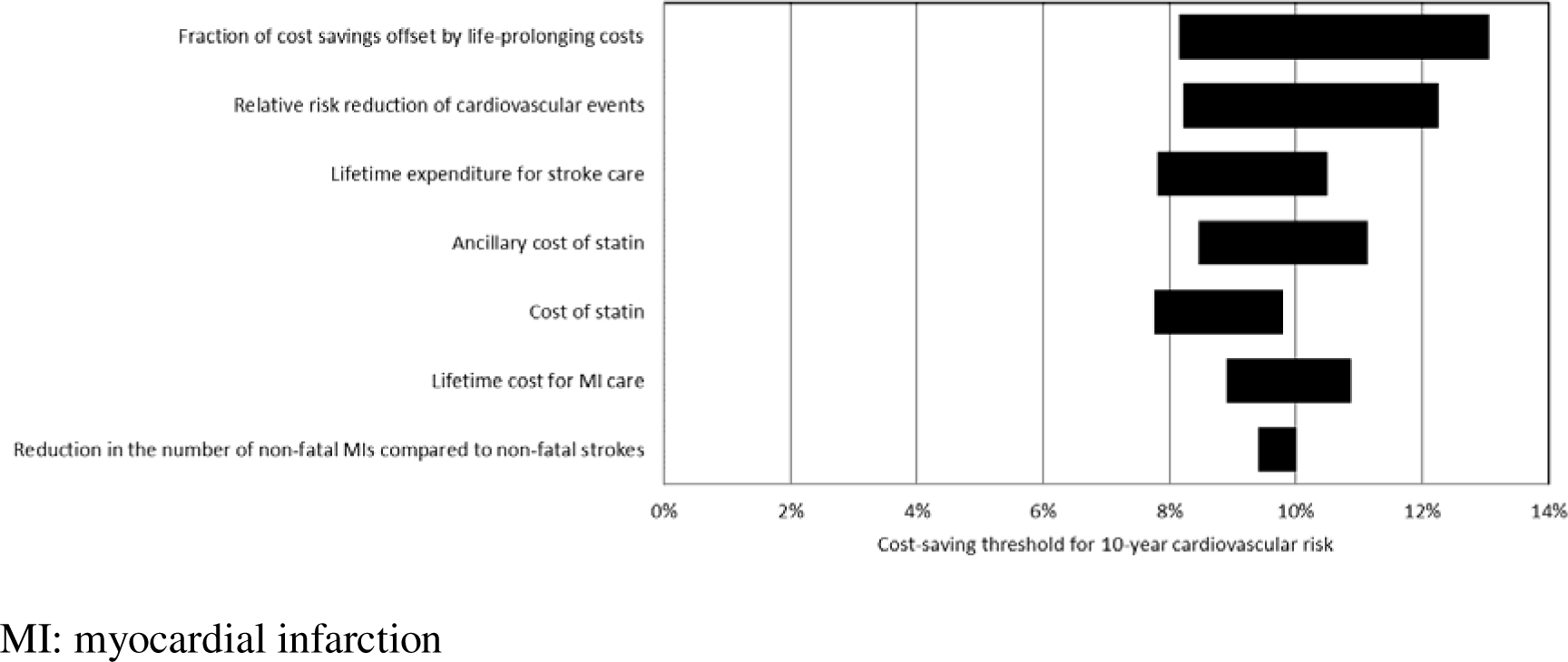
One-way sensitivity analysis of the cost-saving threshold for 10-year cardiovascular risk. Variables are arranged in order of their impact on the 10-year risk threshold.

**Figure 2.**
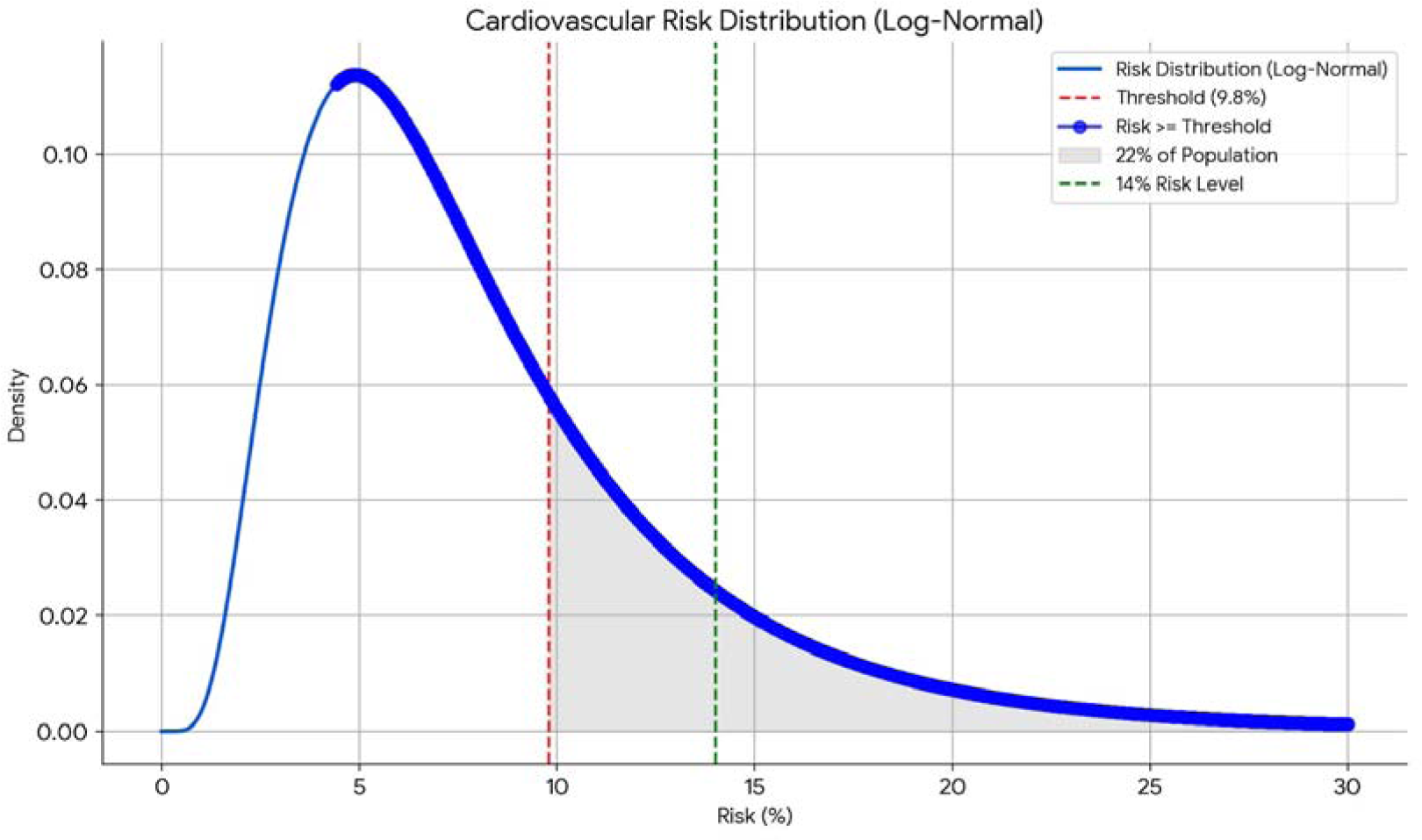
Estimated distribution of 10-year cardiovascular risk in the German population. Dashed lines indicate minimum cost-saving threshold and average risk with a risk above the threshold.

Based on the risk distribution curve, we assumed that around 22% of the adult population in Germany might have a 10-year CVD risk of ≥9.8% (Figure 2). Assuming that the average risk above that threshold ranges between 12% and 15%, the number of avoided CV events over 10 years is estimated to be between 307,049 and 705,537, with a best-case estimate of 475,627, or 47,563 per year (Table 2). The resulting net population savings for sickness funds are approximately €18 billion over 10 years, or €1.8 billion annually (Table 3).

### Sensitivity Analysis

The variable with the greatest impact on the cost-saving threshold for 10-year CV risk was the fraction of cost savings offset by life-prolonging costs, followed by the lifetime cost for stroke care. The one-way sensitivity analysis indicated that the cost-saving threshold for 10-year CV risk consistently remains at or below 13%.

## Discussion

Based on the results of this analysis, the current official threshold for statin prescription in Germany, set by the Joint Federal Committee at a 20% 10-year risk, is too stringent (G-BA, 2020). The risk threshold for statin use determined in this study aligns with the recommendations of the European Society of Cardiology (ESC) for individuals aged 50 to 69 years, which is a 10% CV disease risk (Visseren, 2021). Similar to the ESC recommendations, this risk threshold does not differentiate between primary and secondary prevention. While decisions to initiate statins should prioritize patient health outcomes over cost considerations alone, ensuring that financial incentives do not override clinical judgment, the justification for lowering the threshold relates not only to savings but also to more CV events avoided.

A significant portion of the population could benefit from statin therapy under the new threshold. A population of 13 million sickness fund insurees who would have an indication for statins based on the cost-saving threshold is more than twice as large as the current number of statin users. The uptake rate represents the upper bound of the overuse rate, even in the presence of off-label use (Gandjour, 2018). This indicates that the degree of statin underuse must be larger than the degree of statin overuse in sickness fund insurees.

The rate of underuse is relevant when interpreting the number of avoided CV events and population savings. Although underuse extends above 50% for the newly defined 10-year risk threshold, a portion of CV events is avoided already today by statin use. Hence, the total number of CV events avoided and costs saved reflect the maximum benefit rather than an additional benefit over current practice. Assuming that only one-third of patients are already taking statins above the threshold (based on a total prescription number of about 5 million), we obtain approximately 32,000 CV events avoided per year and savings of €1.2 billion per year (€1.8 billion minus €600 million). Based on the incidence of MIs and ischemic strokes in Germany, statins prescribed above the risk threshold have the potential to reduce this clinical burden by approximately 6%.

The risk threshold for older adults might need adjustment due to different risk-benefit profiles and potential polypharmacy issues. Ensuring that older adults are not overprescribed statins due to competing risks is critical. Although the lifetime costs used in this study were based on a median age of approximately 75 years (Kolominsky-Rabas, 2006), it is prudent to use the threshold cautiously for patients above this age. Despite a shorter remaining life expectancy of stroke and MI survivors above the age of 75, most expenditures for stroke and MI occur in the first year after the incident, which is relatively unaffected by the age of the incident. Based on a registry analysis of stroke patients with a mean age of 76 years from Sweden, mortality after ischemic stroke increases by 6% with each additional year (Sennfält, 2019). Assuming a proportional reduction in expenditure, this would lead to a proportional increase in the cost-saving risk threshold by 0.6% per year. Therefore, the sensitivity of the threshold to the age of stroke or MI incidence is limited. For instance, to increase the 10-year risk threshold from 9.8% to 15.8%, the age of incidence would need to increase from 75 to 85 years.

While it is possible to calculate gender-specific risk thresholds based on gender-specific healthcare savings, this approach may be ethically controversial.

The savings per individual from statin use, as calculated in this analysis, can serve as a benchmark for the upper limit of a reward to users of statins. The annual reward could be as high as €130, which is larger than the cost of the statin itself. Ensuring that patients adhere to their medication to qualify for the reward involves a multifaceted approach combining technology and behavioral strategies. Smart pill bottles provide dose reminders and log each time the bottle is opened. To confirm ingestion, additional methods such as video verification, cholesterol level checks, ingestible sensors, and wearable devices monitoring physiological changes post-ingestion can be employed.

Alternatively, the analysis allows determining the maximum annual costs of statins at which they can be made available without copayments while remaining cost-neutral for sickness fund insurees. This is calculated as the annual cost based on the daily cost of 19 cents plus the annual savings of statins, totaling approximately €202. Currently, a medication can be exempt from copayments if its price is at least 30% below the reference price, which is the maximum amount statutory health insurance covers for certain medication groups.

A cost-effectiveness analysis by Lazar et al. (2011) using U.S. data identified several scenarios where statin use is cost-saving. The scenario closest to this study involves a 10%-20% risk of coronary heart disease (CHD) over 10 years, two risk factors, and a low-density lipoprotein cholesterol level of >100 mg/dL. Notably, the 10-year risk in this scenario refers specifically to CHD, not CV disease as in this paper. This increases the cost-saving treatment threshold because CHD is less expensive than strokes, but additional risk factors included in the scenario increase CV risk, thus lowering the cost-saving treatment threshold.

Previous research in Germany includes a cost-effectiveness analysis that examined the costeffectiveness of statin therapy in primary prevention for different baseline risks (Lauterbach, 2005). The study concluded that statin therapy is cost-effective even at a 5-year risk of 7.5% from the perspective of statutory health insurance. The least favorable cost-effectiveness ratio was found at a 5-year risk of 7.5%, with €26,000 per life year gained for 70-year-old women. From the perspective of social health insurance, pension expenditures significantly increase the cost-effectiveness ratio. However, this study was conducted before generic statins became available in 2003 (Nickolaus, 2003), potentially underestimating the cost-effectiveness of statins due to the drop in prices.

Additionally, a recent cost-effectiveness analysis compared additive lipid-lowering therapies to statin monotherapy for primary and secondary CV prevention in Germany (Michaeli, 2023).

The main limitation of this study is the modeling assumptions, although extensive sensitivity analysis was conducted to test the range of possible values. The study did not use a Markov model, which is typically used for cost-effectiveness modeling and often provides greater precision of estimates. However, Markov models carry assumptions such as a finite set of health states and memoryless transitions occurring at discrete intervals. These assumptions were avoided by using longitudinal cost data from the literature that accounted for the true curvilinear shape of the survival curve in each year post-stroke (Kolominsky-Rabas, 2006). Moreover, the approach taken in this study allows for directly comparing savings, life extension costs, and intervention costs, thus enabling a direct inference of the NNT, which is key to defining the risk threshold indicating savings potential. This also leads to more transparent calculations, which is crucial for convincing policymakers and changing health policy.

Costs of implementation, such as those for public health campaigns, were not considered in the analysis. These may also lead to spillover effects on lower-risk subgroups without cost savings.

The generalizability of this study’s findings to other healthcare systems depends on similarities in population demographics, healthcare infrastructure, and economic conditions. While countries with comparable healthcare systems, cost structures, and guidelines may find the results more applicable, significant differences in medication costs, healthcare policies, insurance models, and epidemiology of CV diseases can affect the relevance and applicability of the study’s conclusions. Therefore, each country should consider local data and context-specific factors to determine the cost-effectiveness and appropriate use of statins for CV prevention within their healthcare system.

For future research, it is recommended to investigate parameters impacting the 10-year risk threshold, as demonstrated in the sensitivity analysis.

## Declarations

Consent for publication: Not applicable
Acknowledgement: None.

## Data Availability

All data produced in the present work are contained in the manuscript

